# Further development of spinal cord retreatment dose estimation: including radiotherapy with protons and light ions

**DOI:** 10.1101/2020.12.20.20248585

**Authors:** Joshua W. Moore, Thomas E. Woolley, John W. Hopewell, Bleddyn Jones

**Author notes:** Correspondence to Prof Bleddyn Jones: Gray Laboratory, Department of Oncology, University of Oxford, Old Road Research Campus Research Building, Roosevelt Drive, Oxford OX3 7DQ, UK.

## Abstract

A new graphical user interface (GUI) was developed to aid in the assessment of changes in the radiation tolerance of spinal cord/similar central nervous system tissues with time between two treatment courses. The GUI allows any combination of photons, protons (or ions) to be used as the initial, or retreatment, course. Allowances for clinical circumstances, of reduced tolerance, can also be made. The radiobiological model was published previously and has been incorporated with additional checks and safety features, to be as conservative as possible. The proton option includes use of a fixed RBE of 1.1 (set as the default), or a variable RBE, the latter depending on the proton linear energy transfer (LET) for organs at risk. This second LET-based approach can also be used for ions, by changing the LET parameters. GUI screenshots are used to show the input and output parameters for clinical situations used in worked examples from previous publications, where the proton and ion treatments required additional ‘longhand’ calculations. The results from the GUI are in agreement with previously published calculations, but the results are now rapidly available without tedious and error-prone manual computations. The software outputs provide a maximum dose limit boundary, which should not be exceeded. Clinicians may also choose a lower number of treatment fractions, whilst using the same dose per fraction (or conversely a lower dose per fraction but with the same number of fractions) in order to achieve the intended clinical benefit. The new GUI will allow rational estimations of time related radiation tolerance changes in the spinal cord and similar central nervous tissues (optic chiasm, brainstem), which can be used to guide the choice of retreatment dose fractionation schedules.

## 1 Introduction

Previous publications have outlined the development of a time-interval dependent model for estimating changes in spinal cord retreatment radiation tolerance [1–4], including the use of a Graphical User Interface (GUI) for photon based retreatment calculations [3,4]. The model incorporated all known radiobiological experimental data sets, as well as a human dose response curve for radiation myelopathy and the time lag before recovery commences. There are also allowances for important clinical factors, which may influence radiation tolerance such as age, previous surgery, and chemotherapy. Although the use of proton therapy was included in the most recent publication [4], with allowances for linear energy transfer (LET) and relative biological effectiveness (RBE), the retreatment dose fractionation estimations required further manual calculations to be carried out independently of the GUI version available at that time [3]. The GUI now provides these solutions, thus obviating the need for such calculations.

The present paper describes a new version of the GUI, which allows retreatment dose fractionation estimations for the spinal cord (and regions of the brain with similar radiation tolerance) for any combination of initial treatment and retreatment using photons, or protons, including the option of two photon courses, or two proton courses. The proton option in any of the treatment combinations may also be changed for ion beam treatments.

## 2 Further Development of the BED calculator

The current GUI has been developed in Java, using the JavaFX library along with Scene Builder to create the actual User Interface (UI). This means that the GUI can be used on any operating system and the file size is extremely low (approximately 100kb), which enables the file to be easily downloaded for clinical use. The new version was designed to have a similar aesthetic to the previous version, originally created in Matlab [3], to allow for a seamless transition to the new calculator. There are small changes regarding notation, therefore a notation popup was added to reduce potential user ambiguity (Appendix 1).

The time-dependent model for photon radiotherapy, developed previously in [3], was used, allowing the same numerical techniques to solve for the appropriate values. To include the use of proton and ion radiotherapy in the GUI, the proton (or ion) doses were converted into photon equivalent doses by using either the conventional standard (fixed) RBE of 1.1 (for protons), or a variable LET-dependent RBE as the dose conversion factor, as developed elsewhere [4], which allow for higher RBE values and thus a lower iso-effective dose. This offers further conservatism. Each of the RBE approached can be applied for each combination of photon and proton (or ion) retreatments, as illustrated in Appendix 2.

The GUI is partitioned into four sections containing each of the potential combinations of treatment sequences mentioned above. Each window runs independently of the others and, consequently, the most recent calculation is stored in its specific window, this enables the user to compare outputs from each treatment combination with minimal effort.

For proton (ion) retreatments, the output of the GUI was changed to provide the number of retreatment fractions for a given proton (ion) retreatment dose per fraction, whereas for photon retreatments the retreatment dose per fraction is the output for a pre-selected number of retreatment fractions. This change is necessary because the retreatment dose per fraction (*d*_ret_) and if used the operative LET must be specified by the user in order to determine the appropriate RBE. Therefore, the only unknown parameter required to estimate the retreatment BED (*BED*_ret_) is the number of retreatment fractions, *n*_ret_. The number of retreatment dose fractions, in clinical practice, can only be a positive integer, but the method used allows for *n*_ret_ to be a non-integer. Thus, a ‘dose adjustment’ feature was implemented to modify *d*_ret_ so that *n*_ret_ rounds to the nearest integer. Using the rounded *n*_ret_ a standard mathematical bisection method [5] was used to solve for small adjustments to *d*_ret_ required for the given *BED*_ret_ value [5]. Further ‘catches’ were developed to inform the user if the suggested dose per fraction changes the RBE value, namely, the RBE output will flash to gain the attention of the user. It is advised that the user first arrives near (within ± 0.5) the desired *n*_ret_ value, then turn on the dose adjustment feature to fine tune the retreatment dose for the integer *n*_ret_. It should be noted that the displayed retreatment dose per fraction is rounded to 2 decimal places in order to be consistent with the photon retreatment windows.

As in the original GUI, various safety procedures have been incorporated into the software. Every input box prevents the input of inappropriate values [3]. If such an inappropriate value was used in any input box, activation of calculation will change the value back to that of the input default. In addition to this, warnings to indicate any initial over-dosage or risks of high probabilities of myelopathy have been introduced. Accompanying these features, by utilising the model developed previously [3], the conservative factor box shifts are also present for proton (or ion) radiotherapy. The most critical of the patient-related inputs (years before retreatment, the conservative factor and retreatment dose per fraction or number of fractions) have been highlighted using borders. These inputs are left empty on launching the application and requires the user to manually set these important parameters. If the user was to leave one of these empty then on starting a calculation, a warning would appear, and the critical empty input box would flash.

## 3 Using the GUI

The first requirement of the user is to choose the appropriate combination of initial and retreatment options. Each possible combination of initial and retreatment options corresponds to one of the tab titles (see supplementary information files to view GUI structure and layout).

Once the appropriate treatment sequence has been selected it is necessary to allocate a percentage risk of myelopathy acceptable on retreatment. The previous default value was set as 1%, but most users preferred 0.1% (the revised default), or a value of 0.1185%, which is the risk associated with the standard assumption that tolerance is associated with a BED value of 100 Gy2 (25 dose fractions of 2 Gy per fraction, total dose 50 Gy). The dose-fractionation details of the initial treatment are then inserted. The time interval between the initial treatment and the proposed retreatment needs to be entered, along with any clinical requirement to change the conservative factor [3]. The proposed number dose fractions for retreatment with photons is then entered, or for all treatments using protons it is necessary to accept one of the two following methods of working:

1. To accept a fixed RBE of 1.1 (the currently accepted standard practice in many centres). This is currently set as the default by the tick in the appropriate box, or can be unticked to:
2. Allow the provision of an appropriate LET value for central nervous tissue, which is converted to an operative RBE value. This also changes the tolerance doses using the equations published previously [4]. This option will provide further protection against any inadvertent over-dosage at low doses per fraction, if RBE values exceed 1.1 in the CNS [6,7].

For ion beam based treatments, it is always necessary to allocate an operative *LET*_*u*_ value (the LET at which RBE is maximal over the entire LET range for any given dose). Recommended values of *LET*_*u*_, published elsewhere [8], are included in the supplementary material.

The choice of an appropriate reference radiation LET (the *LET*_*c*_ value) is also important and should be based on a comparison with megavoltage photons used in the clinic (range 0.2-0.6 keV/*µ*m). The default value is set at 0.22 keV/*µ*m. It should be appreciated that for ortho-voltage x-rays the LET values can significantly exceed 1 keV/*µ*m, especially if poorly filtered, resulting in proton RBE values less than 1 in the mid spread-out Bragg peak (SOBP) [9,10]. The SOBP regions normally have LET values of 1-2 keV/*µ*m, but significantly higher values are found towards the end of the SOBP. Values between 2-10 keV/*µ*m may be found in normal tissues exposed to lower doses outside the Bragg peak region with scanned proton beams [11]. These regions need careful clinical consideration, as suggested previously [4], since the higher LET values increase the RBE.

On completing the input values, the ‘Calculate’ button will become active. Once pressed, the GUI will compute the required solution and will provide one of two outputs, depending on whether retreatment is with photons or protons. In the case of photon retreatments the dose per fraction is displayed, for the requested number of dose fractions. Alternatively, in the case of proton (or ion) retreatment the number of dose fractions is displayed, as a non-integer is displayed if ‘turn off dose adjustment’ is checked. Unchecking the ‘turn off dose adjustment’ will allow the GUI to adjust the dose per fraction, providing a modified dose per fraction size that results in the required full integer fraction number.

The number of fractions proposed for retreatment can be changed and iterations of different dose estimations can be obtained for different conditions, e.g. for alternative treatment plans with different LET values.

When carrying out repeated estimations, it is important to check that the values of the variable parameters remain appropriate. It is recommended that two people with experience in the use of the GUI should check on the parameter entries and the final estimation for clinical use must be interpreted according to the clinical circumstances.

## 4 Results

In order to help new users, window displays are given for various worked examples, largely based on examples used in a previous publication [4]. As a training exercise, users are encouraged to replicate further examples given in the same source.

Various treatment combinations are used in Examples 1-4 (Figures 1-4), and an additional example of a carbon ion retreatment is given in Example 5 (Figure 5). In Example 5 it is necessary to open the fourth (proton and proton option) and then adjust the LET settings.

**Figure 1:**
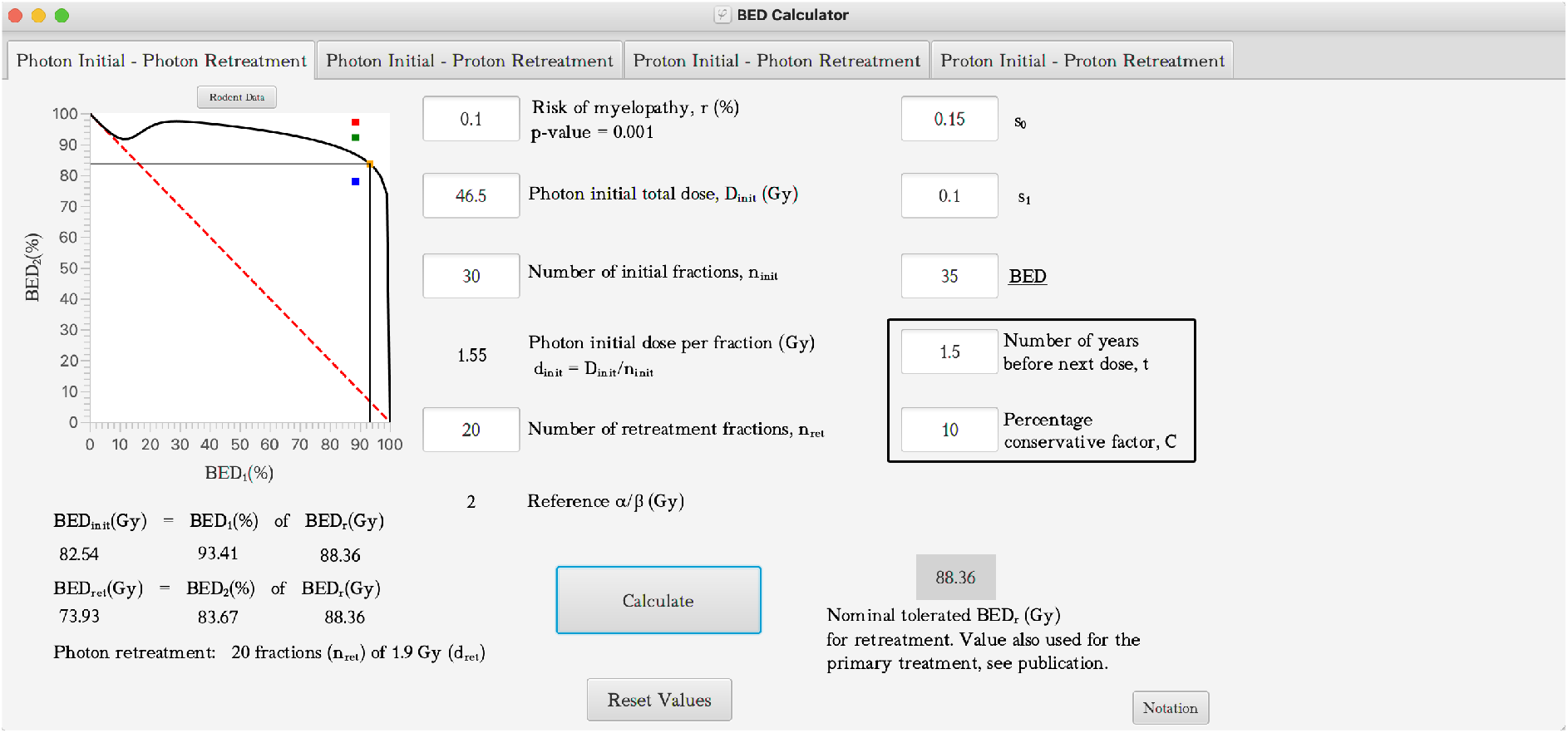
Illustrating the use of the BED calculator via worked example 1 for a Photon-Photon treatment plan. All parameters associated with the time-dependent radiotherapy model (*s*_0_, *s*_1_ and *<u>BED</u>*) were defined previously [3].

### 4.1 Example 1: Initial photon treatment followed by photon retreatment

This is the same example as used with the previously available version of the GUI [3], where the patient received an initial spinal cord dose of 46.5 Gy in 30 fractions. Retreatment for recurrence was required 18 months later. Retreatment was proposed using 20 fractions, with a conservative factor of 10%, due to previous chemotherapy. Figure 1 shows where these inputs have been added as well as the calculated initial dose per fraction (1.55 Gy) and the retreatment dose per fraction of 1.9 Gy, all based on a retreatment myelopathy risk of 0.1% (the BED is 88.36 Gy_2_ after taking into account the additional risk due to chemotherapy). For further explanation of the parameters *s*_0_, *s*_1_ and *<u>BED</u>* see the previous publication [3].

### 4.2 Example 2: Initial photon followed by proton retreatment

The first example of this category uses a known LET allocation while the second example uses a fixed RBE of 1.1 for the same clinical situation. After an initial photon dose of 47.5 Gy in 30 fractions to the spinal cord, a proton retreatment is proposed 18 months later. There were no adverse clinical features so the conservative factor used was 0%. In the first example (Figure 2a) the proton LET was 1.5 keV/*µ*m and the required dose per fraction was entered as 1.6 Gy. The initial output showed a non-integer number of fractions (23.93), so the calculation was repeated with a proton dose of 1.5968 Gy to be given for 24 fractions. It should be noted that in the publication [4], a different approach was used, the number of fractions of 1.6 Gy were rounded down to 23 as a ‘fail safe’.

**Figure 2:**
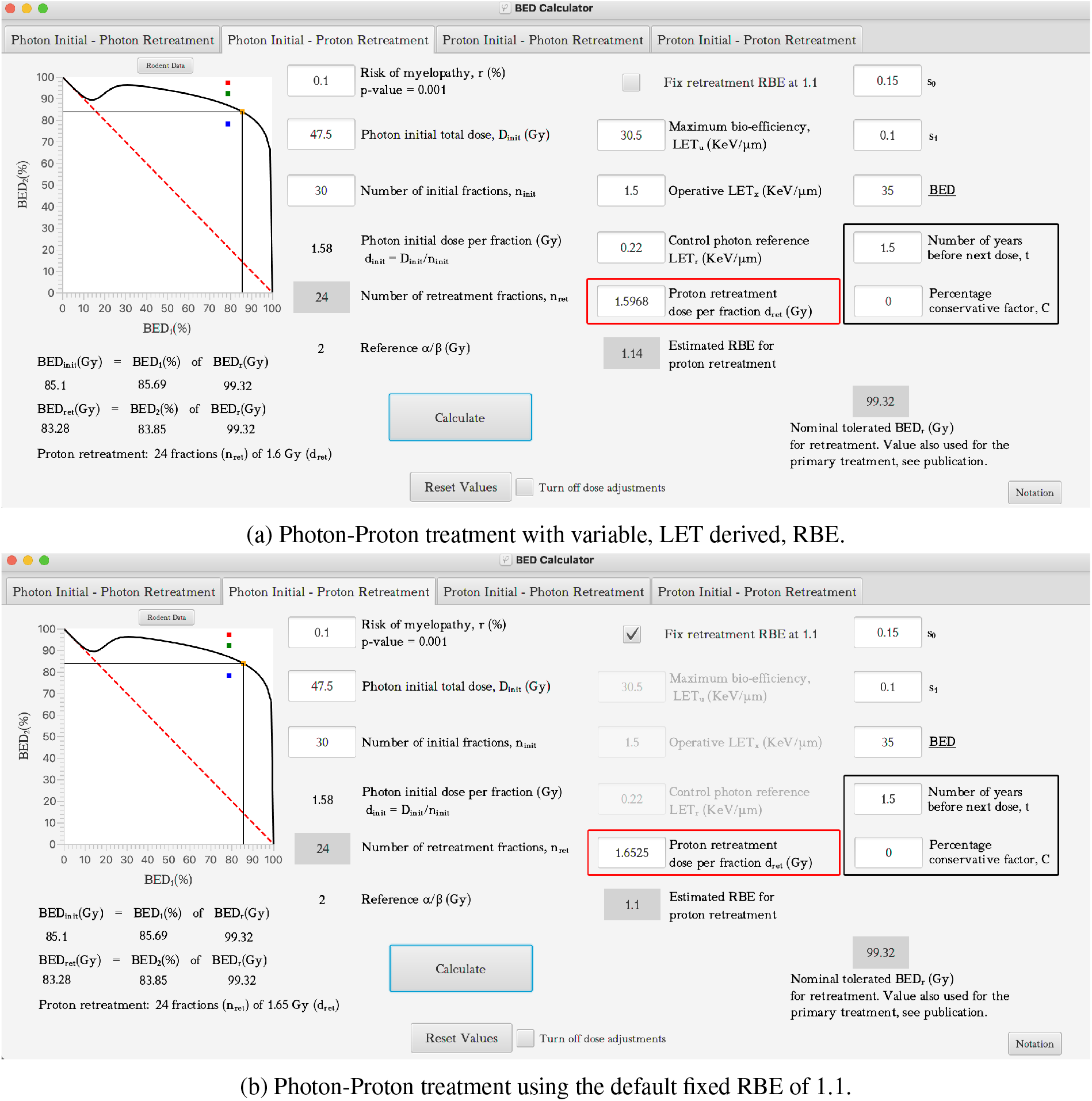
Photon-Proton treatment using the default fixed RBE of 1.1. Figure 2. Illustrating the use of the BED calculator via worked example 2 for a Photon-Proton treatment plan for both variable (a) and fixed (b) RBE. All parameters associated with the time-dependent radiotherapy model (*s*_0_, *s*_1_ and *<u>BED</u>*) were defined previously [3].

In the second example for this treatment combination, the ‘fix retreatment RBE of 1.1’ was left as the default. The output result for a proton retreatment dose per fraction of 1.6525 Gy is 24 fractions. This approach results in a higher total dose being delivered in 24 fractions. The difference total dose delivered was 1.32 Gy, if very accurate doses per fraction were used. Alternatively, the difference in total dose delivered was 1.25 Gy, if the clinically used doses per fraction were rounded to 1.65 and 1.6 Gy, respectively. The smaller dose per fraction of 1.5968 Gy (1.6 Gy) resulted from using an operative RBE of 1.14 as opposed to the fixed value of 1.1. For higher LET values the worked examples in a previous publication [4] should be followed.

### 4.3 Example 3: Initial proton followed by photon retreatment

This example duplicates worked example 6 from previous publication [4] for an initial proton treatment of 39 Gy in 30 fractions to the optic chiasm, with an operative LET of 1.5 keV/*µ*m. Retreatment was required two years after the initial treatment. There is no adverse medical history (conservative factor 0%). The RBE is displayed as 1.15 and for a required 30 photon retreatment fractions, the dose per fraction output is 1.66 Gy (Figure 3).

**Figure 3:**
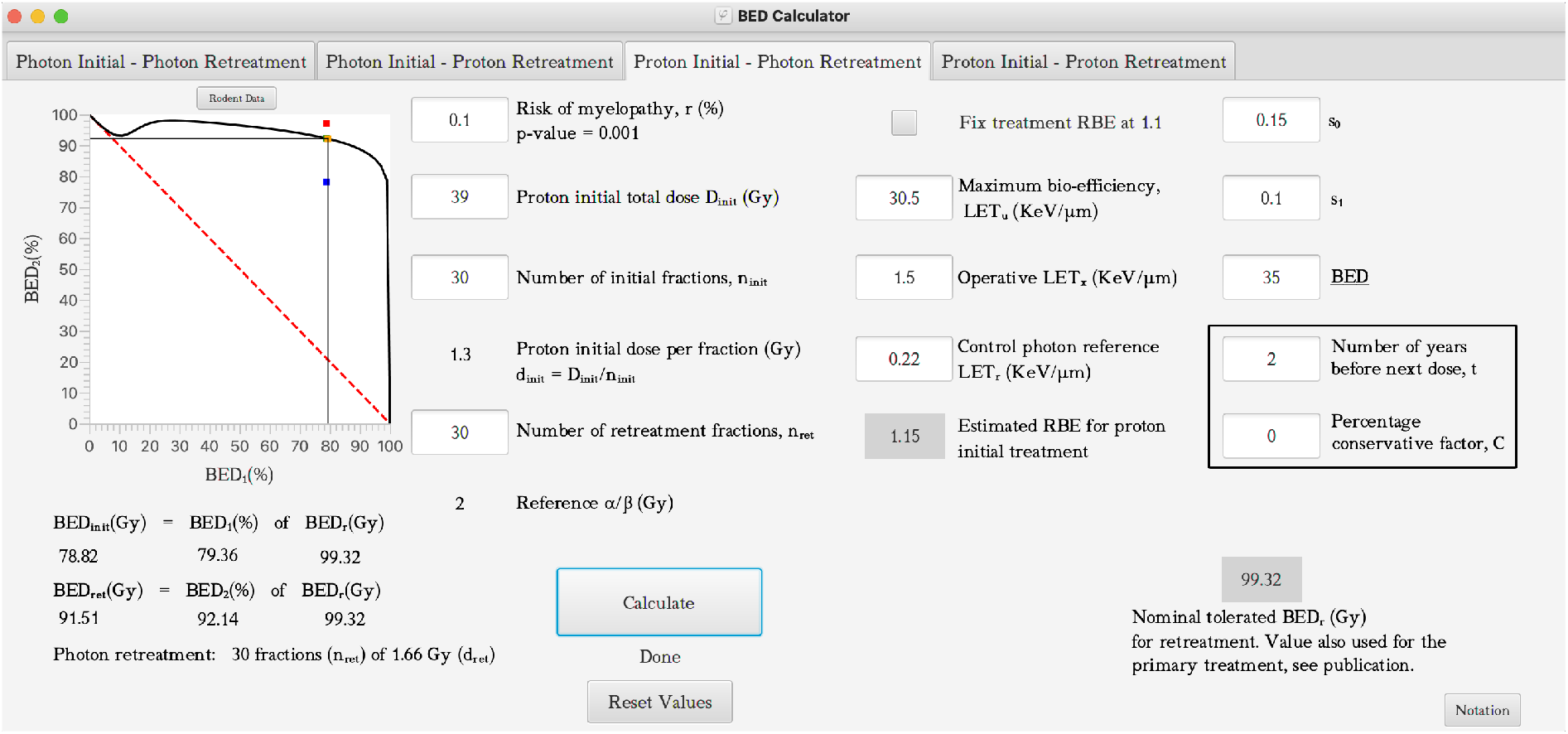
Illustrating the use of the BED calculator via worked example 3 for a Proton-Photon treatment plan. All parameters associated with the time-dependent radiotherapy model (*s*_0_, *s*_1_ and *<u>BED</u>*) were defined previously [3].

### 4.4 Example 4: Initial proton followed by proton retreatment

Here, it is assumed that the first treatment has an LET of 1.3 keV/*µ*m and the retreatment of 1.8keV/*µ*m. The conservative factor is 10% and retreatment is given 2.5 years after the initial treatment of 38.5 Gy in 30 fractions to the spinal cord. The output shows 22 fractions of 1.6747 Gy. In this case, the initial dose per fraction for retreatment used was 1.6 Gy but in order to obtain an integer number of fractions the dose per fraction was changed slightly.

### 4.5 Example 5: Initial carbon ion treatment followed by carbon ion retreatment

In this example two carbon ion treatments were required 2.5 years apart with an allowance for a conservative factor of 10%. The maximum bio-efficiency LET was set at 150 keV/*µ*m for both the initial and retreatment doses. The operative LET was assumed to be 50 and 60 keV/*µ*m for the initial treatment and retreatment, respectively. The initial treatment delivered to a total dose of 10 Gy, in 15 fractions, to the spinal cord. Retreatment was requested using 1.5 Gy dose-fractions. A dose per fraction of 1.5007 Gy was calculated to be consistent with 11 fractions as shown in Figure 5.

**Figure 4:**
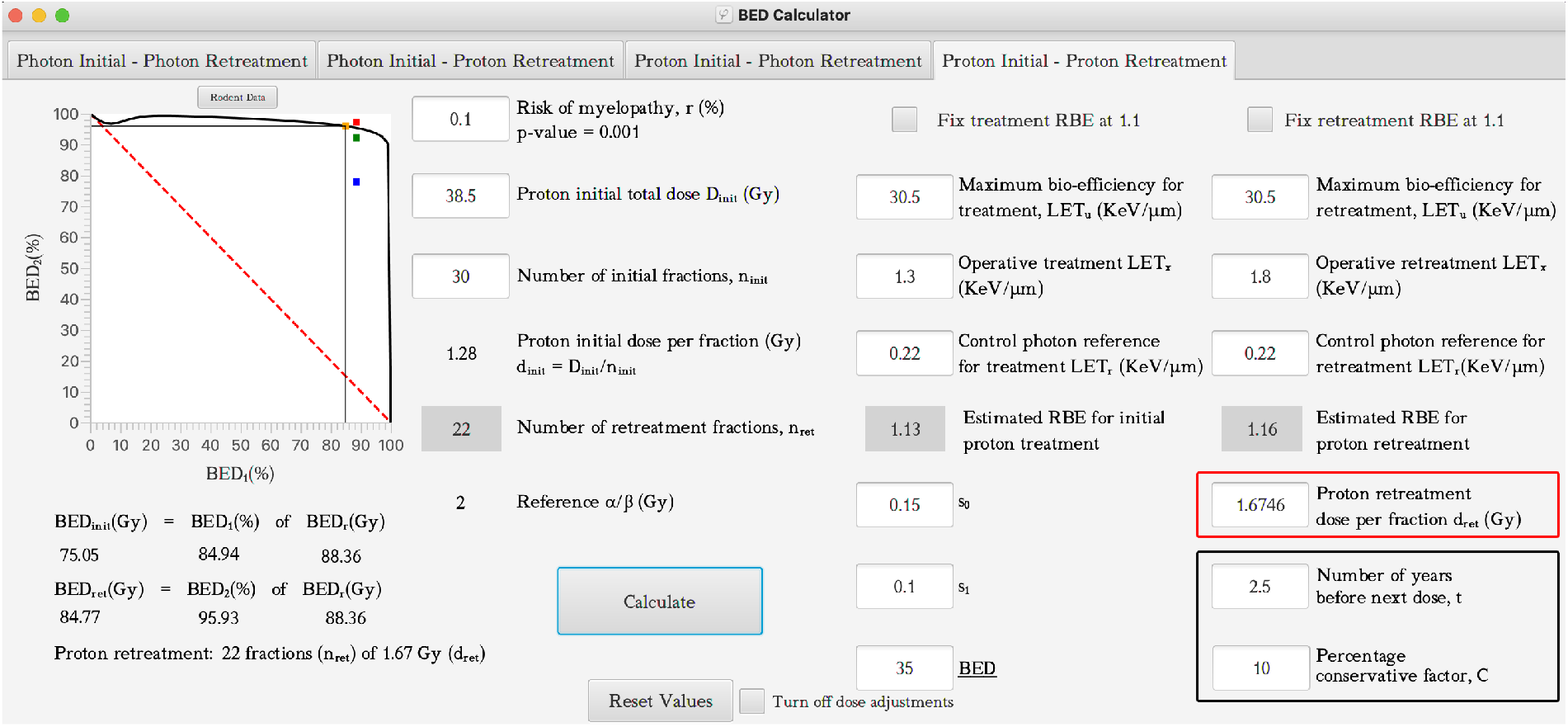
Illustrating the use of the BED calculator via worked example 4 for a Proton-Proton treatment plan, using a LET derived RBE for protons. All parameters associated with the time-dependent radiotherapy model (*s*_0_, *s*_1_ and *<u>BED</u>*) were defined previously [3].

**Figure 5:**
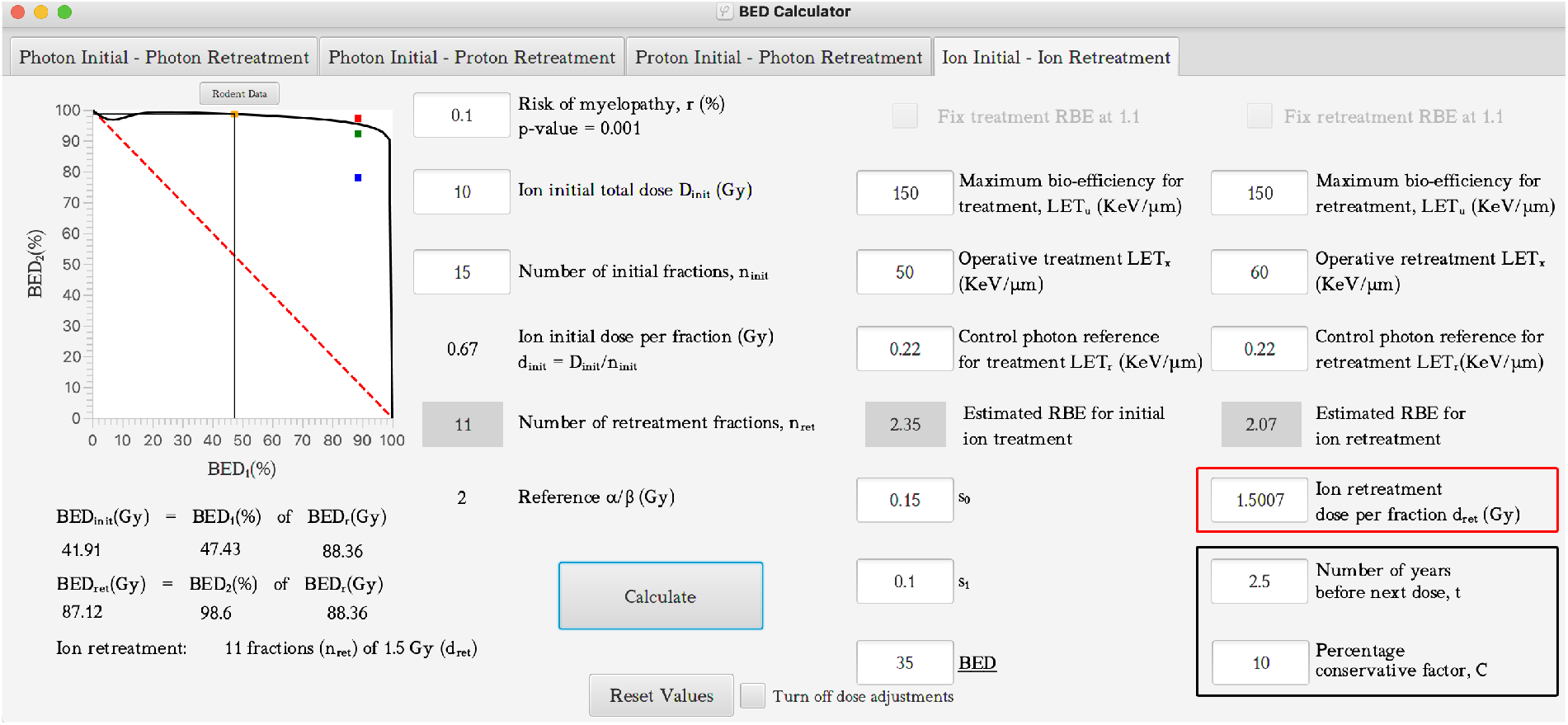
. Illustrating the use of the BED calculator via worked example 5 for a carbon ion – carbon ion treatment plan. All parameters associated with the time-dependent radiotherapy model (*s*_0_, *s*_1_ and *<u>BED</u>*) were defined previously [3].

**Figure 6:**
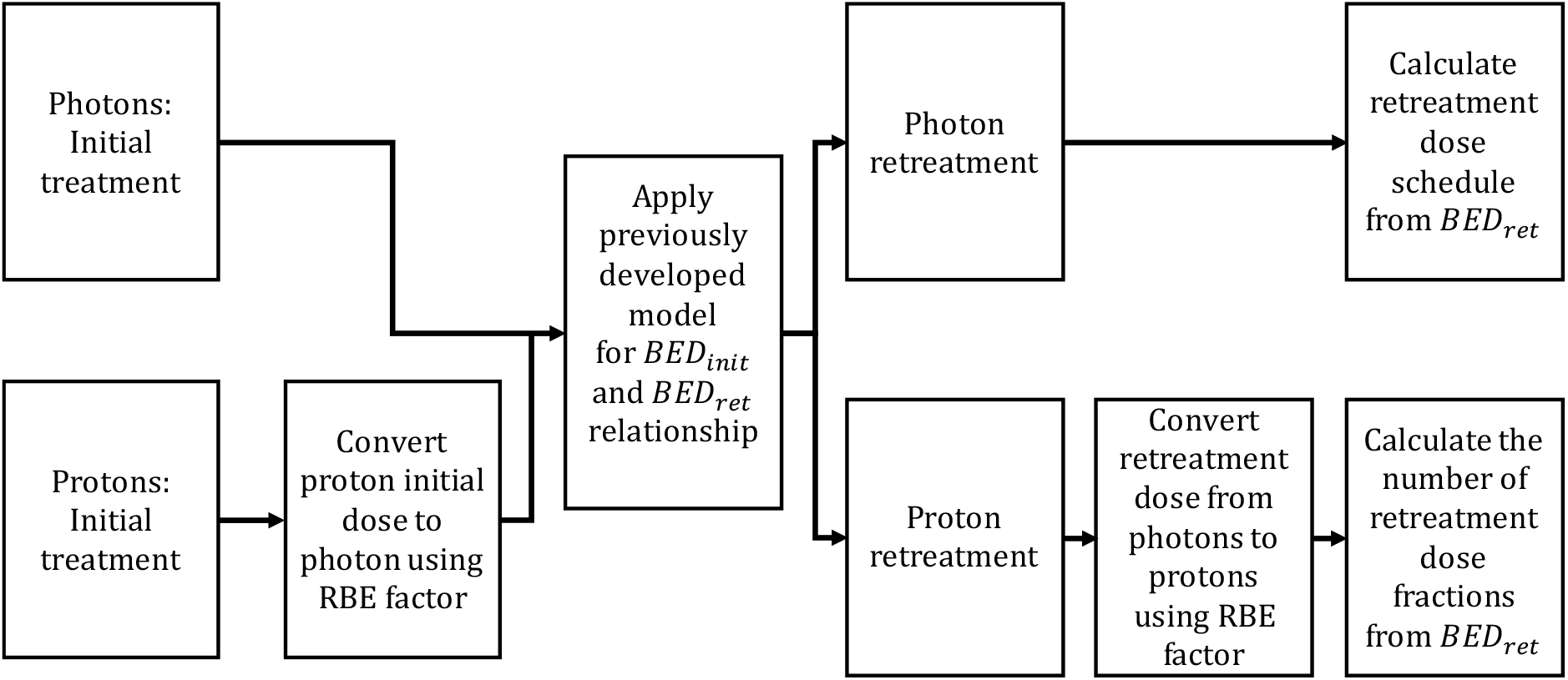
Computational pipeline for the BED calculation for both Photon and Proton treatments.

## 5 Discussion

The new GUI described above provides a practical platform for obtaining retreatment dose-fractionation estimations for the spinal cord and other areas of the central nervous system with similar radiation tolerance, such as the brainstem and optic chiasm regions. It removes the need to do any additional manual calculations, which have many separate steps and pitfalls, especially when the added difficulties presented by increasing LET and RBE are involved.

The system aims to provide a safe <u>upper limit</u> to the retreatment dose per fraction for a given number of fractions, but continues to need considerable clinical input to define the acceptable risk level for a given retreatment situation, and to determine the degree of conservative factor allocation (a BED reduction allocation according to clinical circumstances, as discussed previously [3,4]). Further, clinical judgement may be required to operate in a ‘fail safe’ way, for example by using a lower number of retreatment fractions than suggested, if there is any doubt. The upper limit provided by the model is more appropriate for radical retreatment situations. For palliative situations there is considerable leeway for reductions in dose per fraction or fraction number in order to deliver a schedule that has a high probability of achieving the intended effect, such as relief of pain or bleeding, which require a lower BED. Such situations are often delivered using fraction sizes greater than 2 Gy and even single treatment sessions in some countries.

The system presented has used only one LET-RBE model, but this is based on results from extensive biological data sets, with wide ranges of radiobiological parameters; however, many others are available [12]. Since the GUI is open source such models could potentially be included in any further extensions. It is further proposed that the use of the selected dose-fractionation, on retreatment, when using the default proton RBE of 1.1 should be compared with that obtained using the LET bases RBE model if the relevant tissue LET is known, or can be assumed on the basis of previously published studies. Some clinicians may prefer to adopt an intermediate treatment value between the estimates provided by each of these two approaches.

As in most clinical modelling systems, important caveats are required:

1. The results are dependent on the validity of the linear quadratic model of radiation effect and on all the assumptions made.
2. The retreatment dose-fractionation estimates represent a boundary condition, which should not be exceeded.
3. It cannot be over-emphasised that the output estimations act as a guide to patient management and the final dose per fraction, or number of fractions estimated, may be reduced according to the preferences of the treating clinician.
4. The risks of retreatment must be accepted by the responsible clinician, who should be familiar with the underlying radiobiological principles, and in all cases full informed consent should be obtained after discussions with each individual patient.

If used with care, the GUI should allow clinicians and physicists to estimate safe retreatment doses when radiation treatments with photons and protons (or ion beams) are given as separate treatment courses.

## Supporting information

BED Calculator

BED Calculator - readme

BED Calculator Guidelines

BED Calculator Annotated Diagrams

BED Calculator Getting Started

BED Calculator Reference LET Values

## Data Availability

The data used in this study is sourced from prior works which are referenced throughout the manuscript.

## Acknowledgements

JWM acknowledges the support of Cardiff University via funding from the Cardiff Undergraduate Research Opportunities Programme (CUROP) 2019 and Knowledge Economy Skills Scholarships (KESS2), KESS2 is a pan-Wales higher-level skills initiative led by Bangor University on behalf of the Higher Education sector in Wales. It is part-funded by the Welsh Government’s European Social Fund (ESF).

## Appendix 1

The revised Graphical User Interface (GUI) described in this manuscript can be downloaded as zip file from the Supplementary data associated with this manuscript. This contains a Java exe file and the associated font file. A Read Me file sets the terms for use of the GUI along with guidelines.

## Appendix 2

Flow diagram showing pathways for the calculation processes used in the revised GUI. The initial treatment is inputted as the total dose and number of dose fractions. Proton (ion) doses are converted to equivalent photon dose using an RBE factor, so that the initial treatment can be assigned a *BED*_*init*_ for the level of risk considered acceptable on retreatment, including conservatism thought appropriate on clinical grounds. The *BED*_*ret*_ is then converted back to an acceptable photon treatment directly or via an RBE factor if retreatment is to be with protons (ions).

