## Supplementary material for "Further development of spinal cord retreatment dose estimation: including radiotherapy with protons and light ions": BED Calculator - readme

A supplementary GUI for the paper titled 'Further development of spinal cord re-treatment dose estimation: including radiotherapy with protons and light ions'.

BED CALCULATOR

For details behind the calculator's operation and usage please see:

Further development of spinal cord re-treatment dose estimation: including radiotherapy with protons and light ions. J. W. Moore, T. E. Woolley, B .Jones, J .W . Hopewell xxxx

Changes in the retreatment radiation tolerance of the spinal cord with time after the initial treatment. T. E. Woolley, J. Belmonte-Beitia, G. F. Calvo, J. W. Hopewell, E. A. Gaffney and B. Jones. Int J Radiai Biol xx, xxx-xxx, 2108

Spinal cord re-treatments using photon and proton based radiotherapy: LQ-derived tolerance doses. B .Jones, J .W . Hopewell. Euro J Medical Phys

TERMS AND CONDITIONS

By accessing and using the GUI you accept the following conditions:

Any clinical decisions taken following the use of the GUI must be the responsibility of the local clinical specialists of the user.

The GUI must be used in conjunction with the advice in the original published papers.

Previous GUI Versions (Original GUI)

Updated versions of the original GUI will also been made available using the following supplementary link,

http://thomaswoolley.co.uk/codes

Current GUI Versions

Updated versions of the current GUI will also been made available using the following supplementary link,

https://github.com/joshwillmoore1/BED-Calculator

Any computational issues related to the use of the GUI can be reported on the appropriate GitHub page under "issues". Note Thomas E. Woolley and Joshua W. Moore are mathematicians and are unable to supply any help on any particular medical application. These should be directed to the local clinical specialist of the user.

GETTING STARTED

Download the zip file which contains the GUI titled "BED_Calculator.jar".
Check if you have Java installed on your computer - this can be done by opening the console (Windows+R then type cmd) or the equivalent for your operating system. In the console type "java -version" and press enter. If java is installed a version number will be displayed, if not then download the environment from this link - https://java.com/en/download/manual.jsp.
Once the Java environment is installed on your computer, extract the contents of the zip file to a location of your choice.
Before starting the calculator, install the provide font file "quivira.tff". Simply double-click this file to install it.
Finally, to start the calculator, double-click the "BED_Calculator.jar" file.
If the GUI fails to open, please see the getting started file provide.
KNOWN ISSUES

Some subscript notation may not be displayed. If this is the case, please install the provide font in the font file and restart the GUI. If the B̅E̅D̅ notation is not being displayed and you're using a windows OS, try using the MAC version of the GUI.

Windows users - IF DOUBLE-CLICK DOES NOT START APPLICATION then please check the Java version installed on the PC is the same version in the registry. This can be done by pressing windows key+R and searching for "regedit". Then navigate down the 'CLASSES_ROOT' file to find the 'jarfile' folder. Continue to open this file until the file 'command' is reached. The text in this file should then be edited to the version that is installed in either the 'program files' or 'program files x86'. Locate the bin file and open it (e.g. C:\Program Files\Java\jdk1.8.0_191\bin). Copy this address and insert it into the registry 'command' file text in the appropriate position ( e.g. "C:\Program Files\Java\jdk1.8.0_191\bin\javaw.exe" -jar "%1" %* ). Press ok and close registry. The app should now open on double-click.
