## Supplementary material for "Further development of spinal cord retreatment dose estimation: including radiotherapy with protons and light ions": BED Calculator Guidelines

Biomed. Phys. Eng. Express xx, xxx – xxx, 20xx

BED CALCULATOR

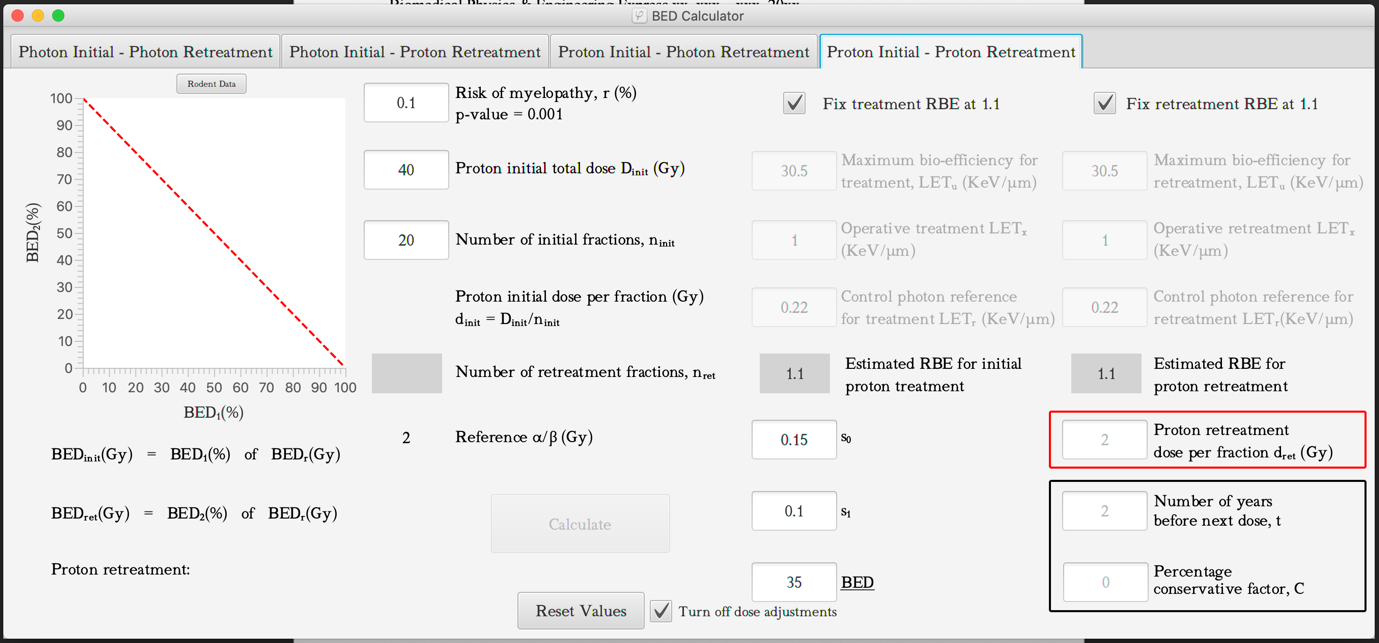

The Graphical User Interface (GUI) described in this manuscript can be downloaded as an executable in the zip file provided entitled ‘BED Calculator’.

Updated versions of the GUI will also be made available using the following supplementary link: -

<https://github.com/joshwillmoore1/BED-Calculator>

Any technical computational issues related to the use of the GUI can be report directly to. Please note Thomas E. Woolley and Joshua Moore are mathematicians and are unable to supply any help on any particular medical application. These should be directed to the local clinical specialist of the user.

By accessing and installing the BED calculator, or Graphic user interface (GUI), to a personal computer the user is also agreeing to accept the following conditions:

**TERMS AND CONDITIONS**

1) Any clinical decisions taken following the use of the GUI must be the responsibility of the local clinical specialists of the user.

2) The GUI must be used in conjunction with the advice in the original published paper.
