## Supplementary material for "Further development of spinal cord retreatment dose estimation: including radiotherapy with protons and light ions": BED Calculator Annotated Diagrams

Biomed. Phys. Eng. Express xx, xxx – xxx, 20xx

This document contains example screenshots of the GUI. Figure 1 is an example of the GUI layout for a proton retreatment while Figure 2 is an example of the GUI for a photon retreatment. It must be emphasized that there are distinctions between the two. In particular, there are many differences between the inputs and outputs. The annotated figures are placed here to aid the user and should be used in conjunction with the main text.

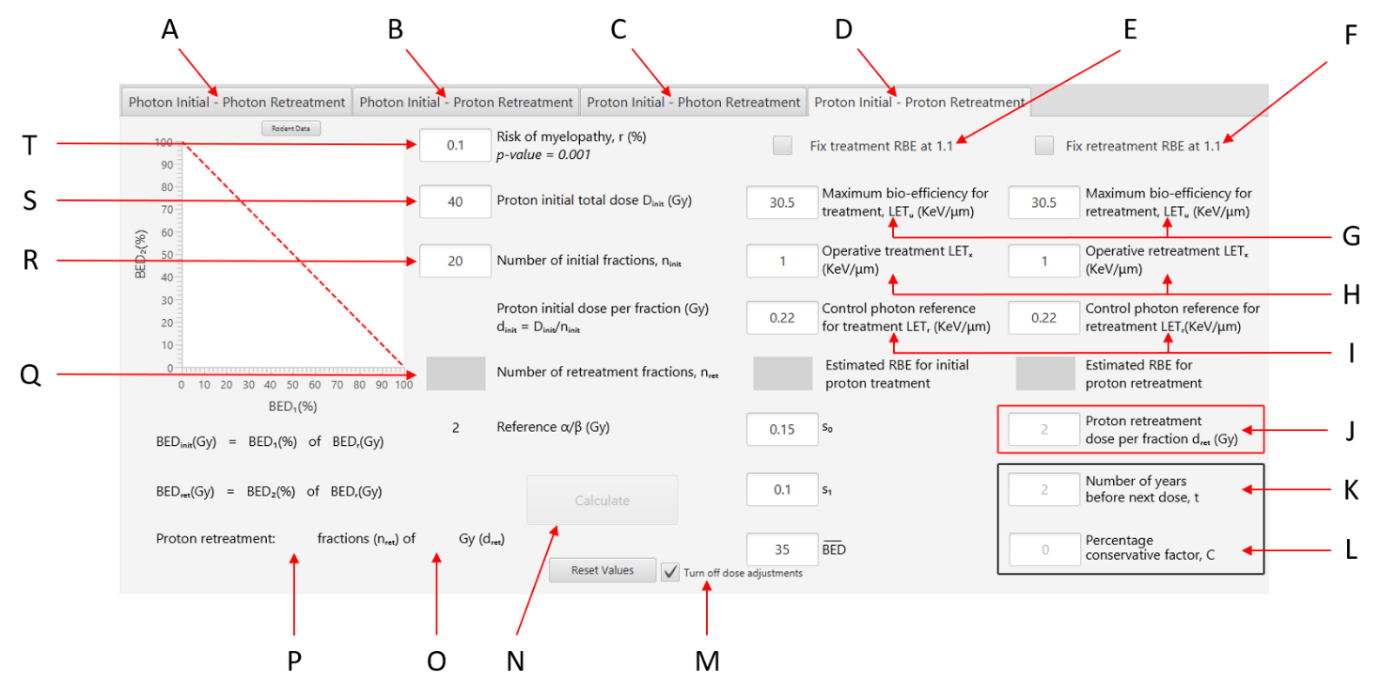

**Figure 1. Annotated screenshot of the GUI with initial proton treatment and proton retreatment window selected and default values. A-D are the tab buttons that are used to select the corresponding combination of proton-photon treatments. E-F are checkboxes that are by default ticked, when the GUI to set for use with a fixed RBE value of 1.1 for the initial proton treatment and proton retreatment, respectively and must be unticked to allow input into. G the** $\mathbf{LET}_{\mathbf{u}}$ **input boxes for both initial treatment and retreatment and H the** $\mathbf{LET}_{\mathbf{x}}$ **input boxes for both initial treatment and retreatment. I refers to the** $\mathbf{LET}_{\mathbf{r}}$ **input boxes for both initial treatment and retreatment. J is the input box for retreatment proton dose. K is the input box time-elapsed between initial treatment and retreatment. L is input box for the conservative factor. M corresponds to the checkbox that turns off the dose adjustments. N is the calculate button (initially inactive, but becomes active once all inputs are added). O refers to where the retreatment doses are displayed. P refers to where the number of retreatment fractions are displayed. Q displays the number of retreatment fractions for the proton retreatments and is an input box for the number of retreatment fractions for photon retreatments. R-S refer to the initial dose and T is the input box for the acceptable risk of myelopathy.**

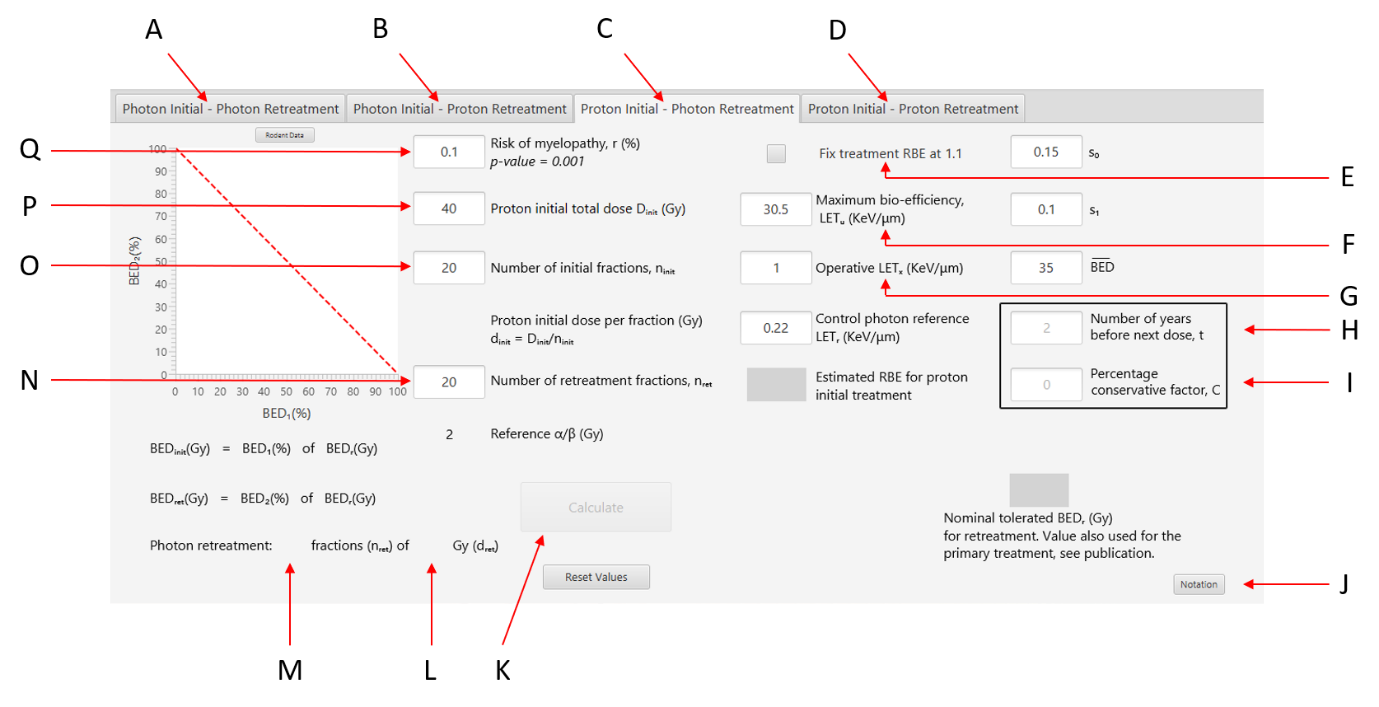

**Figure 2. Annotated screenshot of the GUI with initial proton treatment and photon retreatment window selected and default values. A-D are the tab buttons that are used to select the corresponding combination of proton-photon treatments. E is the checkbox that is ticked when the fix default RBE value of 1.1 for the initial proton dose is set. F refers to the** $\mathbf{LET}_{\mathbf{u}}$ **input boxes for initial treatment. G refers to the** $\mathbf{LET}_{\mathbf{x}}$ **input boxes for initial treatment. H is the input box time-elapsed between initial treatment and retreatment. I is input box for the conservative factor. J corresponds to the notation button that opens the window of definitions. K is the calculate button (initially inactive, but becomes active once all inputs are added). L refers to where the retreatment doses are displayed. M refers to where the number of retreatment fractions are displayed. N is the input box for number of retreatment fractions. P-O refer to the initial dose and Q is the input box for the acceptable risk of myelopathy.**
