## Supplementary material for "Further development of spinal cord retreatment dose estimation: including radiotherapy with protons and light ions": BED Calculator Getting Started

Biomed. Phys. Eng. Express xx, xxx – xxx, 20xx

The GUI was designed to be highly accessible with minimum software requirements. Java Runtime Environment (JRE) is all that is needed to run the GUI and most computers have this already downloaded. Once JRE is installed, the GUI can be downloaded from the supplementary materials and doubled-clicked to start. Here, we outline the steps to be taken to get the BED Calculator running any machine:

1. Install the font provide (Optional). The “quivira.tff” font file is included with the calculator. Double-click this file and follow the instructions to install the font.
2. Open the BED Calculator. If you already have JRE install on your computer, you should be able to double-click the “BED_Calculator.jar” file to start up the software and you should be greeted with a splash page. If this does not open the calculator, then continue to the next step.
3. We first check that JRE is installed. Navigate to your terminal/command prompt and type “java -version” and press enter. If no version number is displayed, then go to <https://java.com/en/download/manual.jsp> to download JRE (this is open source, free software). Once downloaded try opening the “BED_Calculator.jar” file again.
4. If a java version number is displayed in the terminal/command prompt and the “BED_Calculator.jar” file is still not opening, then there may be a security feature preventing the use of apps from unidentified developers (a common problem for Mac users). To open the calculator, navigate to system preferences, then go to security and privacy. Under the “general” tab, press allow unidentified app to open, this may require your computer password. After this step, the calculator should open.

For more troubleshooting advice, please see the READ ME document provided.
