## Supplementary material for "Further development of spinal cord retreatment dose estimation: including radiotherapy with protons and light ions": BED Calculator Reference LET Values

**Biomed. Phys. Eng. Express xx, xxx – xxx, 20xx**

Point estimates of *LET_U_* obtained by linear regression techniques on multiple ion beam data sets which provide RBE values based on cell survival changes with LET, using a range of radiation doses, are given in the table:

| ***Ion*** | ***LET_U_ (keV.μm^-1^)*** |
| --- | --- |
| proton | 30.4 |
| helium | 120.1 |
| carbon | 157.1 |

For other ion species the LET_U_ can be estimated from the Z (nuclear charge values) using the empirical relationship

$$\boldsymbol{LET}_{\boldsymbol{U}}\boldsymbol{=30.4+}\boldsymbol{108.4}/\boldsymbol{0.61}\mathbf{Exp}\left[ \boldsymbol{1-0.61}\sqrt{\boldsymbol{(Z-1)}} \right]$$

Source: Jones B. and Hill M. A. Physical Characteristics at the Turnover-points of Relative Biological Effect (RBE) with Linear Energy Transfer (LET). Biomed. Phys. Eng. Express 6 (2020) 055001 https://doi.org/10.1088/2057-1976/ab9e13
